# Use of commercially available natural language processing software to identify bleeding from the medical record

**DOI:** 10.1101/2021.11.19.21266586

**Authors:** Andrew L. Walker, Cheri Watson, Ryan Butcher, Zameer Abedin, Mark Yandell, Rashmee U. Shah

## Abstract

**Background:** Real-world evidence derived from the electronic medical record (EMR) is increasingly prevalent. How best to ascertain cardiovascular outcomes from EMRs is unknown. We sought to validate a commercially available natural language processing (NLP) software to extract bleeding events.

**Methods:** We included patients with atrial fibrillation and cancer seen at our cancer center from 1/1/2016 to 12/31/2019. A query set based on SNOMED CT expressions was created to represent bleeding from 11 different organ systems. We ran the query against the clinical notes and randomly selected a sample of notes for physician validation. The primary outcome was the positive predictive value (PPV) of the software to identify bleeding events stratified by organ system.

**Results:** We included 1370 patients with mean age 72 years old (SD 1.5) and 35% female. We processed 66,130 notes; the NLP software identified 6522 notes including 654 unique patients with possible bleeding events. Among 1269 randomly selected notes, the PPV of the software ranged from 0.921 for neurologic bleeds to 0.571 for OB/GYN bleeds. Patterns related to false positive bleeding events identified by the software included historic bleeds, hypothetical bleeds, missed negatives, and word errors.

**Conclusions:** NLP may provide an alternative for population-level screening for bleeding outcomes in cardiovascular studies. Human validation is still needed, but an NLP-driven screening approach may improve efficiency.

## Introduction

Electronic medical records (EMR) are increasingly prevalent and contain a multitude of data that can be analyzed for research purposes. Clinical events can be captured from the EMR using International Classification of Diseases (ICD) codes, which allows for pragmatic, large-scale research studies. However, ICD codes in EMR can be inaccurate, leading to misclassification^1,2^.

Natural language processing (NLP) is a technology that mines the unstructured narrative and provides an alternative to the ICD code-driven approach to identify events from EMR. Commercial software for healthcare NLP is increasingly available, but studies to validate accuracy are limited. The goal of this project was to measure the performance of CliniThink, a commercially available NLP software, to identify location-specific bleeding events among patients with atrial fibrillation (AF) and cancer. Bleeding is a common adverse event in cardiovascular disease, and a key endpoint in clinical trials and quality improvement initiatives. Plus, the use electronic medical records (EMRs) for clinical investigation and regulatory processes is increasing.^3–5^ A NLP solution to find bleeding events across large populations could improve efficiency and reduce the cost of executing these studies.

## Methods

We performed a retrospective analysis of AF patients treated at the Huntsman Cancer Institute at University of Utah Health from 1/1/2016 to 12/31/2019. The cohort was constructed using the health system’s enterprise data warehouse (EDW), which integrates data from different sources for both operations and research. AF patients were selected using our previously developed NLP algorithm^6^. The resulting patients were then merged with a registry of cancer patients maintained by the Huntsman Cancer Institute to arrive at the patient population with both conditions. For each patient, we defined the index date (“day 0”) as the date of first visit to the cancer hospital for patients with a prior AF ICD code, or as the date of the first AF ICD code at the cancer hospital for patients without a prior AF ICD code. We chose this approach using the logic that clinical decisions (e.g., anticoagulation treatment) are made at the time of joint AF/cancer diagnoses. Demographics were extracted from the EDW, drug exposures (apixaban, dabigatran, edoxaban, rivaroxaban, and warfarin) were derived from medication order and reconciliation tables, and comorbid conditions were derived by grouping ICD 10 codes into clinically meaningful groups.

We used the NLP software to identify bleeding in the population. The software requires the user to provide a “query” set based on SNOMED CT (Systematized Nomenclature of Medicine Clinical Terminology, https://www.snomed.org/) expressions in the form of a text document. An example SNOMED CT expression is as follows: 243796009|Situation with explicit context|:{408731000|Temporal context|=410512000|Current or specified|,246090004|Associated finding|=249366005|Bleeding from nose|,408732007|Subject relationship context|=410604004|Subject of record|,408729009|Finding context|=410515003|Known present|}. This expression indicates epistaxis, or nosebleed, occurring during the encounter and to the patient. We developed a bleeding specific query using 226 terms, aggregated into 11 anatomic categories: neurologic; ophthalmologic; ear, nose, and throat (ENT); pulmonary; gastrointestinal (GI); genitourinary (GU); obstetrician/gynecology (OBGYN); cardiac; orthopedic; retroperitoneal hematoma; and general category with other types of bleeds. To limit reporting small sizes, we report the 7 most frequently occurring anatomic categories in this report. The Python script and resulting query text file can be found here: https://github.com/rashmeeushah/nlp_bleeding.

The query was executed on notes written by physicians, advance practice clinicians, and pharmacists. To identify these notes from the large corpus of EMR notes, we used a simple regular expression matching approach. Clinical notes in our EDW contain a header that includes “author type” and we limited these types to terms specific for the clinicians of interest. We validated the positive predictive value (PPV) of the query at the level of the note. Notes were selected randomly for manual review by a physician to determine if bleeding was absent or present based on the text, taking a sensitive approach to adjudication. In other words, if bleeding text was written in the note, we erred on the side of clinically significant. We chose this approach because, at this stage, an implemented system to identify bleeds would probably involve human adjudication and verification, which would exclude the false positive results. We then did a manual error analysis to identify patterns related to false positives. This retrospective study was exempted by the University of Utah Health Institutional Review Board with a waiver of informed consent.

## Results

The population of AF and cancer patients included 1370 patients, with mean age 72 years old (standard deviation 1.5 years). Among these patients, 35% were female, 92% were Caucasian, and 3.6% were Hispanic. Less than half, 44%, of patients were treated with an oral anticoagulant at the time of the index visit. Other characteristics are found in Table 1.

**Table 1.** Baseline characteristics of 1,370 patients with atrial fibrillation and cancer.

| **Characteristic, n (%) unless otherwise indicated** | **Not reviewed (n=908)** | **Reviewed\* (n=462)** | **P-Value** |
| --- | --- | --- | --- |
| Age, mean years (SD) | 73.5 (10.2) | 70.4 (10.8) | <0.001 |
| Female | 328 (36.1) | 156 (33.8) | 0.422 |
| White | 830 (91.4) | 421 (91.1) | 0.850 |
| Hispanic | 36 (4.0) | 14 (3.0) | 0.354 |
| Atrial fibrillation present prior to initial cancer visit | 432 (47.6) | 216 (46.8) | 0.817 |
| Cancer Type |  |  | 0.015 |
| Neurologic | 38 (4.3) | 21 (4.6) |  |
| Breast | 85 (9.6) | 23 (5.0) |  |
| Gastrointestinal | 158 (17.8) | 81 (17.7) |  |
| Genitourinary | 185 (20.9) | 91 (19.9) |  |
| Gynecologic | 29 (3.3) | 30 (6.6) |  |
| Head and neck | 39 (4.4) | 14 (3.1) |  |
| Hematologic | 138 (15.6) | 93 (20.4) |  |
| Lung | 85 (9.6) | 39 (8.5) |  |
| Other | 130(14.3) | 65 (14.1) |  |
| Anticoagulation ordered or indicated in medication reconciliation | 405 (44.6) | 202 (43.7) | 0.801 |
| Comorbid Conditions |  |  |  |
| Congestive heart failure | 247 (27.2) | 163 (35.3) | 0.002 |
| Coronary artery disease | 381 (42.0) | 225 (48.7) | 0.020 |
| Cerebrovascular disease | 163 (18.0) | 123 (26.6) | <0.001 |
| Chronic kidney disease | 218 (24.0) | 149 (32.3) | 0.001 |
| Chronic lung disease | 268 (29.5) | 163 (35.3) | 0.035 |
| Hypertension | 660 (72.7) | 373 (80.7) | 0.001 |
| Diabetes | 376 (41.4) | 242 (52.4) | <0.001 |
| *\*Reviewed indicates that at least one note from 462 patients was reviewed by a physician to validate a bleeding event.* |  |  |  |

The initial corpus included 134,270 notes for 1370 unique patients. Limiting notes by author type resulted in 66,130 notes including 1,363 unique patients. This corpus was processed in the NLP software (with a runtime of 2 to 3 hours), which identified 6522 notes including 654 unique patients with possible bleeding events. Among these notes, 1269 randomly selected notes for 462 patients were reviewed by a physician. The PPV of the software ranged from 0.921 for neurologic bleeds to 0.571 for OB/GYN bleeds (Table 2). Among randomly selected false positives for review, patterns related to false positive bleeding events identified by the software included historic bleeds (44%), hypothetical bleeds (29%), missed negatives (13%), and word errors (5%). An example of a common hypothetical bleed included notes with transfusion threshold instructions in the assessment and plan and notes where clinicians mentioned the risk of bleeding when outlining a procedure.

**Table 2.** **Positive predictive value for bleeding events identified by natural language processing (NLP) software in clinical notes**

| **Anatomic Category** | **True bleed/NLP identified bleed** | **PPV** |
| --- | --- | --- |
| Neurological | 129/140 | 0.921 |
| Ophthalmologic | 19/21 | 0.905 |
| Pulmonary | 51/59 | 0.864 |
| ENT | 69/84 | 0.821 |
| Genitourinary | 119/146 | 0.815 |
| Gastrointestinal | 193/264 | 0.731 |
| OBGYN | 20/35 | 0.571 |

## Discussion

This work demonstrates the potential for NLP software to screen for clinically meaningful events across large populations using unstructured narrative from the EMR. If implemented, the number of notes that require review could decrease by >90%. An NLP approach would reduce the need for an ICD-driven approach, which lacks both accuracy and clinical granularity. For example, classifying a gastrointestinal bleeding event using ICD codes omits key details, such as intensive care unit admission, blood transfusion, or resultant discontinuation of oral anticoagulation. A streamlined, NLP-driven approach could enable more efficient chart review to identify events and extract salient details.

Areas for ongoing improvement remain. First, the algorithm still requires human review since context cannot easily be resolved from the clinical narrative. Our error review demonstrates issues related to temporal context. SNOMED CT expressions include many components, and over time, these components can be further specified to improve NLP accuracy. For example, we used the “current or specified” temporal context; adding “recent” to temporal context could increase sensitivity. Second, “computable definitions” for outcomes are needed in order for an NLP-driven approach to replace current methods. Current bleeding definitions such as the International Society on Thrombosis and Haemostasis^7^ and Bleeding Academic Research Consortium^8^ are not computable. Eventually, these definitions could be based on SNOMED CT expressions. In the short term, however, an NLP-driven screening process with subsequent adjudication against existing definitions is a reasonable solution.

These results have limitations. First, we studied a limited population of AF and cancer patients. Second, we estimated performance at the level of the note; NLP accuracy at the visit level—including all notes for a single hospitalization, for example— will be needed for future applications. Third, we used data from a single health system. Curating the corpus and the style of text varies between health systems and performance will vary. Finally, to ensure unbiased results, performance should be validated in larger populations according to race/ethnicity, sex, and age.

## Conclusions

Our results support the possibility that healthcare-specific NLP could facilitate rapid, high quality, clinical research and quality improvement across large populations. This approach could reduce chart review burden and enable outcome ascertainment from documentation generated as part of the clinical workflow, and thus enable larger, more generalizable pragmatic and observational studies.

## Data Availability

Protected patient data are not available for sharing. The code to generate the SNOMED CT expressions for the NLP software are publicly available on GitHub, as referenced in the manuscript.

## References

1. Shah, R. U. et al. Impact of Different Electronic Cohort Definitions to Identify Patients With Atrial Fibrillation From the Electronic Medical Record. J. Am. Heart Assoc. 9, e014527 (2020).

2. Faridi, K. F. et al. Comparability of Event Adjudication Versus Administrative Billing Claims for Outcome Ascertainment in the DAPT Study. Circ. Cardiovasc. Qual. Outcomes 14, e006589 (2021).

3. Office of the Commissioner. Real-World Evidence. U.S. Food and Drug Administration https://www.fda.gov/science-research/science-and-research-special-topics/real-world-evidence (2020).

4. Mori, M. et al. The Promise of Big Data and Digital Solutions in Building a Cardiovascular Learning System: Opportunities and Barriers. Methodist Debakey Cardiovasc. J. 16, 212–219 (2020).

5. Khera, R. et al. RESEARCH PROTOCOL: Large-scale evidence generation and evaluation across a network of databases for type 2 diabetes mellitus. bioRxiv (2021) doi:10.1101/2021.09.27.21264139.

6. Shah, R. U. et al. Development of a Portable Tool to Identify Patients With Atrial Fibrillation Using Clinical Notes From the Electronic Medical Record. Circ. Cardiovasc. Qual. Outcomes 13, e006516 (2020).

7. Kaatz, S., Ahmad, D., Spyropoulos, A. C., Schulman, S. & the Subcommittee on Control of, A. Definition of clinically relevant non-major bleeding in studies of anticoagulants in atrial fibrillation and venous thromboembolic disease in non-surgical patients: communication from the SSC of the ISTH. J. Thromb. Haemost. 13, 2119–2126 (2015).

8. Kikkert, W. J. et al. The Prognostic Value of Bleeding Academic Research Consortium (BARC)-Defined Bleeding Complications in ST-Segment Elevation Myocardial Infarction: A Comparison With the TIMI (Thrombolysis In Myocardial Infarction), GUSTO (Global Utilization of Streptokina. J. Am. Coll. Cardiol. 63, 1866–1875 (2014).

